# Angiotensin-converting enzyme inhibition and Alzheimer’s disease: Findings from proteome-wide Mendelian randomization and a large prospective cohort study

**DOI:** 10.1101/2024.09.09.24313366

**Authors:** Peiyuan Huang, Haibo Tang, Zeying Feng, Evie Stergiakouli, Tom R Gaunt, Jie Zheng, Yao Lu

## Abstract

**Background:** We aimed to provide clinically translatable insights for drug discovery, repurposing, and vigilance for preventing Alzheimer’s disease (AD) by integrating genetic and “real-world” drug use data.

**Methods:** Proteome-wide Mendelian randomization (MR) analysis was conducted to identify plasma proteins causally related to AD risk using the largest summary-level data to date, followed by colocalization and multi-omic validation analyses (on the gene expression, alternative splicing, and DNA methylation levels in blood and brain, respectively) to prioritize potential druggable targets. We also replicated our MR findings using additional genetic data and, where appropriate, performed multivariable MR for the prioritized findings. Conventional observational analysis using the data from UK Biobank, a large prospective cohort, was performed to provide further clinical implications for our genetic findings.

**Results:** MR analysis identified 15 plasma proteins with putative causal effects on AD risk. Of them, inhibition of angiotensin-converting enzyme (ACE) was found to increase the risk of AD (OR 1.10, 95% CI 1.08-1.14), which was likely independent of blood pressure as suggested by multivariable MR. Observational analysis in UK Biobank showed a higher incidence (HR 1.24, 95% CI 1.01-1.52) of AD among regular users of ACE inhibitors (ACEI), compared with the counterpart angiotensin receptor blocker users.

**Conclusions:** In addition to expanding the understanding of druggable targets for AD prevention, our findings highlighted the potential risk of AD associated with the use of ACEIs, a widely prescribed antihypertensive medication, suggesting the need for caution in clinical practice and further research on the effect of antihypertensives on neurodegenerative diseases.

## INTRODUCTION

Alzheimer’s disease (AD) is one of the most prevalent neurodegenerative disorders, characterized by irreversible neuronal death and progressive cognitive impairment. As the leading cause of dementia, AD affects approximately 50 million individuals worldwide and poses a significant medical and social burden ^1^. With an aging population, the global incidence of dementia is expected to increase to 150 million by 2050 ^2^. Despite extensive research, the pathogenesis of AD remains elusive, and few effective preventive or therapeutic strategies have been established ^3, 4^. Therefore, there is an urgent need to develop novel approaches to prevent and treat AD.

The human proteome provides a valuable resource for gaining insights into the pathogenesis of AD and identifying potential biomarkers and druggable targets. Our previous study demonstrated the translational value of drug target discovery for complex diseases using proteome-wide Mendelian randomization (MR) ^5^. Since then, several proteome-wide MR studies of blood and/or brain have identified various proteins as potential drug targets for AD ^6–9^. However, these findings were obtained and validated only using genetic tools and lacked observational evidence from “real-world” drug use data, thus limiting their reliability.

Aiming to reveal the pathogenesis of AD and facilitate discovery, repurposing, and vigilance of drugs for AD prevention, we first conducted a proteome-wide MR analysis, followed by sensitivity and replication analyses as well as multi-omic validation. For any targets identified by this process, we integrated observational evidence based on UK Biobank data on drug use to further validate them and explore their clinical implications. An overview of the study design is shown in **Figure 1**. By incorporating “real-world” drug use data into omics-based approaches, we can link omics-based research findings with clinical practice, fostering a deeper understanding of the underlying mechanisms of AD, facilitating the development of more effective preventive measures, or identifying existing drugs that may decrease or increase the risk of AD.

**Figure 1.**
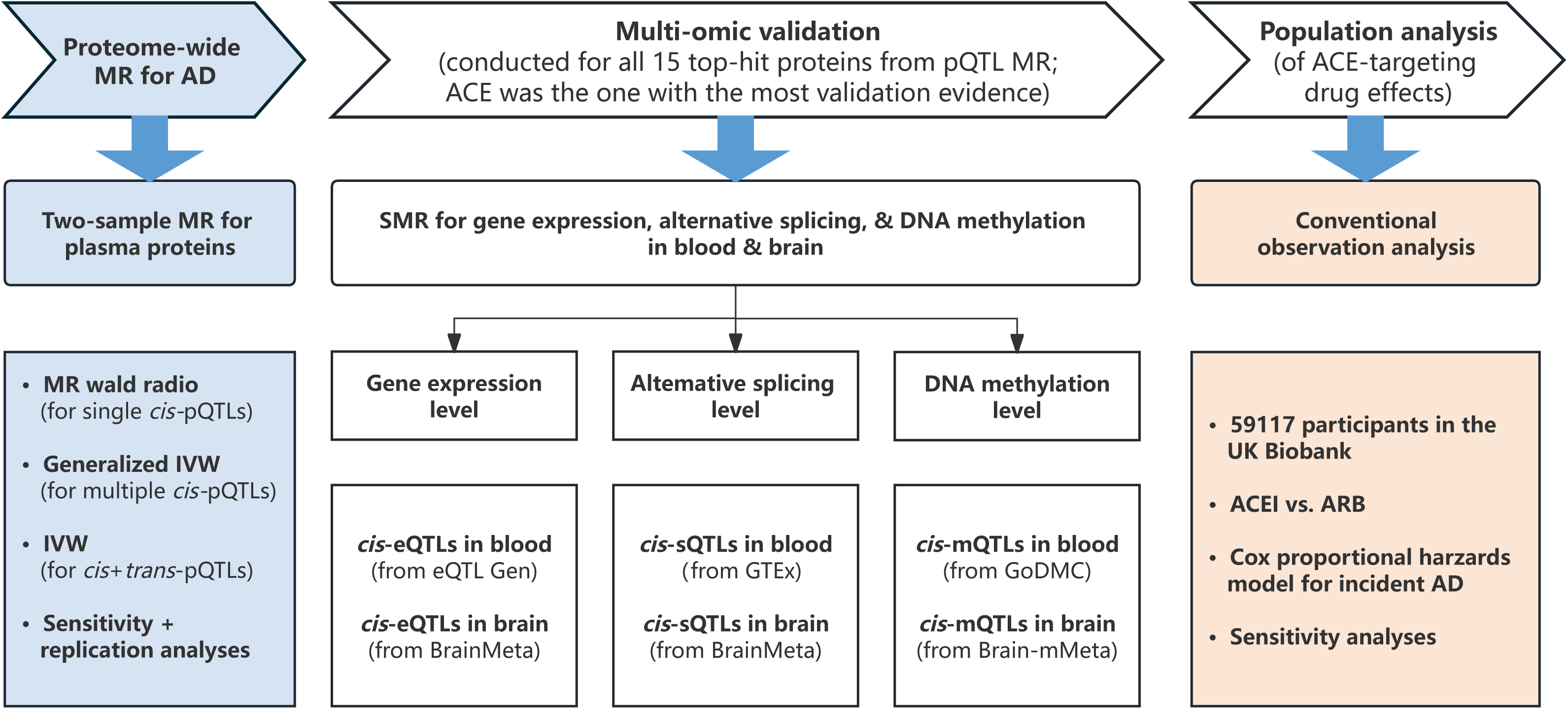
Overview of the study design. MR, Mendelian randomization. AD, Alzheimer’s disease. pQTL, protein quantitative trait locus. IVW, inverse variance-weighted method. SMR, summary-data-based Mendelian randomization. eQTL, expression quantitative trait locus. sQTL, splicing quantitative trait locus. mQTL, methylation quantitative trait locus. DNA, deoxyribonucleic acid. GTEx, Genotype-Tissue Expression. GoDMC, Genetics of DNA Methylation Consortium. ACE, angiotensin-converting enzyme. ACEI, angiotensin-converting enzyme inhibitor. ARB, angiotensin receptor blocker.

## METHODS

### Mendelian randomization

#### Genetic associations with plasma proteins

We used protein quantitative trait loci (pQTL) instrument data from our previous study by Zheng et al. ^5^, which integrated five published pQTL studies and identified 1,064 tier 1 (i.e., with the highest relative level of reliability) pQTL instruments for 955 proteins.^5^ Tier 1 pQTL instruments were further categorized into three groups: (1) *cis*-pQTLs (including single-*cis*-pQTLs [with only one *cis*-pQTL available for each protein] and multiple*-cis*-pQTLs [with more than one *cis-*pQTLs available for each protein]), (2) *cis + trans*-pQTLs, and (3) *trans*-pQTLs. As *trans*-acting pQTLs may operate via indirect mechanisms and are therefore more likely to be pleiotropic ^10^, we only focused on tier 1 single-*cis*-, multiple*-cis*-, and *cis + trans*-pQTLs in this study.

#### Genetic associations with Alzheimer’s disease

Outcome genome-wide association study (GWAS) data were from the latest and largest published AD GWAS by Bellenguez et al. ^11^, including 111,326 clinically diagnosed/“proxy” AD cases and 677,663 controls of European ancestry, from the following consortia/datasets: EADB, GR@ACE, EADI, GERAD/PERADES, DemGene, Bonn, the Rotterdam study, the CCHS study, NxC, and the UK Biobank ^12^. In the UK Biobank, individuals who did not report dementia or any family history of dementia were used as controls, and the analysis included 2,447 diagnosed cases, 46,828 proxy cases of dementia, and 338,440 controls ^11^. There was no sample overlap between the pQTL studies and the AD GWAS.

#### Genetic associations with gene expression, gene expression, alternative splicing, and DNA methylation

Blood *cis-*expression quantitative trait loci (*cis*-eQTL) data were obtained from the eQTLGen Consortium (https://www.eqtlgen.org/), which incorporates 37 datasets to identify the downstream consequences of trait-related genetic variants, with a total of 31,684 individuals ^13^. Brain *cis*-eQTL data were accessed from a recent eQTL mapping analysis by Qi et al ^14^. using RNA sequencing data of brain cortex (N = 2,865) with genome-wide single nucleotide polymorphism (SNP) data. Blood *cis-*splicing quantitative trait loci (*cis*-sQTL) data were obtained from the Genotype-Tissue Expression (GTEx, https://gtexportal.org/) Analysis V8 (N = 670), assessing the association between genetic variants and intron excisions using the LeafCutter approach ^15^. Brain cortex *cis*-sQTL data were also accessed from Qi et al.’s study ^14^, which used an sQTL mapping method called testing for heterogeneity between isoform-eQTL effects (THISTLE) and identified 12,794 genes with *cis*-sQTLs at *P* < 5 × 10^-8^. Blood *cis-*methylation quantitative trait loci (*cis*-mQTL) data were extracted from the Genetics of DNA Methylation Consortium (GoDMC, http://mqtldb.godmc.org.uk/), which identified genetic variants associated with DNA methylation (DNAm) at 420,509 cytosine-phosphate-guanine (CpG) sites in blood among 27,750 individuals of European ancestry. Brain *cis*-mQTL data were obtained from the meta-analysis by Qi et al. (N = 526 to 1194) ^16^.

#### Genetic association data for replication

In addition to the combination of exposure and outcome GWAS data in our MR main analysis (i.e., Zheng et al. – Bellenguez et al.), replication analysis was performed using the data from different pQTL studies and AD GWASes. For the exposure (protein abundance) GWAS, we used the data from a recent pQTL study by Gudjonsson et al ^17^. This study examined 4,035 independent associations between genetic variants and 2,091 serum proteins in 5,368 individuals aged between 66 and 96 years from the AGES-Reykjavik Study ^18^. pQTL full summary data were available for all of our top-hit proteins. For the outcome (Alzheimer’s disease), we used the data from three GWASes. The first one, by Schwartzentruber et al. ^19^, included a total of 75,024 cases (including those with diagnosed AD and who reported a family history of dementia) and 397,844 controls from a previous GWAS ^20^ and the UK Biobank ^12^. The second one, by Kunkle et al. ^20^, was a subset of Schwartzentruber et al.’s GWAS samples, which included 21,982 cases with diagnosed AD and 41,944 controls. The third one, by Wightman et al. ^21^, was a subset of a large GWAS meta-analysis excluding 23andMe data, including 86,531 cases and 676,386 controls. We selected these three GWASes because Schwartzentruber et al.’s and Wightman et al.’s studies were two of the largest and latest AD GWASes following Bellenguez et al.’s study, and Kunkle et al.’s study only recruited diagnosed AD cases, whose results were thus more clinically relevant.

#### Genetic associations with blood pressure

In follow-up analyses, we conducted multivariable MR to adjust for blood pressure. To avoid potential collider bias caused by adjustment for body size in the GWAS ^22^, genetic association data for systolic (SBP) and diastolic blood pressure (DBP) were obtained from the UK Biobank GWASes (SBP GWAS ID: ukb-b-20175; DBP GWAS ID: ukb-b-7992) of ∼436,420 males and females of European ancestry, accessed through the IEU OpenGWAS Project (http://gwas.mrcieu.ac.uk/) ^23^.

### Conventional observational study

#### Study population

The UK Biobank is a prospective cohort study recruiting over 500,000 million middle-aged participants at 22 assessment centers across the UK between 2006 and 2010. When recruited, participants provided blood, urine, and saliva samples at the nearest assessment center with their consent, as well as detailed information about sociodemographic, lifestyle, health-related factors, environment, and medical history via touchscreen and face-to-face interviews. Physical measurements were taken, including height, weight, and blood pressure. Follow-up assessments were conducted using routinely available national datasets. To avoid potential confounding by indications, we restricted our study to participants with either angiotensin-converting enzyme inhibitor (ACEI) or angiotensin receptor blocker (ARB) medication records. Individuals were further excluded if they met any of the following criteria at baseline: 1) with a diagnosis of AD before baseline [International Classification of Diseases 10 (ICD-10) codes: F00 and G30]; 2) with a family history of AD.

#### Measurement of medication use

According to the Anatomical Therapeutic Chemical (ATC) classification ^24^, ACEIs and ARBs were defined with the code C09AA and C09CA, respectively. Combining the medication records at baseline in UK Biobank, participants who had been using the drugs (ACEIs or ARBs) most days of the week for the last four weeks, were classified as regular users of ACEIs or ARBs.

#### Ascertainment of Alzheimer’s disease

AD cases were identified by the diagnosis records with ICD-10 code F00 and G30 ^25^, followed up to 30 September 2021. The date of diagnosis was set as the date of the first AD diagnosis record. Person-time was taken from the date of baseline and censored at the date of AD diagnosis, date of death, or 30 September 2021, whichever occurred earliest.

#### Assessment of covariates

At baseline, participants provided demographic and lifestyle behaviors information, including age at recruitment, sex, ethnicity, smoking status, alcohol consumption frequency, income score (UKB Data Field 26411, with higher scores indicating higher income levels), education score (UKB Data Field 26414; with higher scores indicating higher education levels), and comorbidities. Body mass index (BMI; in kg/m^2^) was calculated by weight (in kg) divided by squared height (in m). The comorbidities were identified by ICD-10 at baseline, including hypertension, diabetes, hyperlipidemia, traumatic brain injury, cerebrovascular diseases, and nervous system diseases (ICD-10 codes: I60-I69). In addition, we also calculated the number of long-term conditions as covariates. Detailed definitions of long-term conditions were described elsewhere ^26^.

### Statistical analyses

#### Proteome-wide Mendelian randomization

For single-*cis*-pQTLs, we performed a standard two-sample MR analysis for each protein with the Wald ratio method ^27, 28^. For multiple*-cis*-pQTLs, MR estimates may be sensitive to the particular choice of pQTLs if only the most strongly associated SNPs within each genomic region are used as instruments ^5^. It has been suggested that more precise causal estimates can be obtained using multiple genetic variants from a single gene region, even if the variants are correlated ^29, 30^. Therefore, we used multiple conditionally independent *cis-* pQTLs for this MR analysis. Specifically, we conducted linkage disequilibrium (LD) clumping with a relaxed threshold of r^2^ < 0.6 to avoid highly LD-correlated SNPs; the resulting single-*cis*-pQTLs (i.e., multiple*-cis*-pQTLs with highly LD-correlated SNPs removed by clumping) were picked back to the single-*cis* analysis above; for the rest multiple*-cis*-pQTLs, we used a generalized inverse variance-weighted (IVW) ^31^ method for each protein considering the LD pattern between the multiple-*cis* SNPs to estimate the MR effects, where the pairwise LD correlation (r^2^) were obtained from the 1000 Genomes European ancestry reference samples. For *cis + trans*-pQTLs, a standard two-sample MR analysis was performed for each protein, with r^2^ < 0.001 as the threshold for LD-clumping and standard IVW as the MR method ^28, 32^. To correct for multiple testing, the Bonferroni method was utilized in the analysis for each of the above three types of pQTLs.

#### Colocalization

Results that passed the multiple-testing threshold in the above MR analysis were further evaluated using the Bayesian colocalization analysis to estimate the posterior probability (PP) of each genomic locus containing a single variant affecting both the protein abundance and AD risk ^33^. Colocalization assesses whether two associated signals are consistent with a common causal SNP. Default prior probabilities were used (*P*_1_ = 1 × 10^−4^, *P*_2_ = 1 × 10^−4^, *P*_12_ = 1 × 10^−5^, where *P*_1_ is the probability that a given SNP is associated with AD, *P*_2_ is the probability that a given SNP is a pQTL, and *P*_12_ is the probability that a given SNP is both associated with AD and is a pQTL). We applied a PP threshold of >80% for the hypothesis that there is a shared causal SNP for both traits as sufficient evidence for colocalization.

In cases where pQTL data lacked sufficient SNP coverage (e.g., without publicly available full summary data), we instead conducted the “LD check” analysis ^5^ for the sentinel variant for each protein against the 30 strongest SNPs in the region associated with AD as an approximate colocalization analysis. r^2^ > 0.8 between the sentinel variant and any of the 30 SNPs with the strongest association with AD was used as evidence for approximate colocalization. For our MR top findings, we treated colocalized findings (PP > 80%) as “Colocalized” and LD-checked findings (r^2^ > 0.8) as “LD-checked”; other findings that did not pass the colocalization or LD check analyses were annotated as “Not colocalized”.

The presence of multiple conditionally distinct association signals within the same genomic region can influence the performance of colocalization analysis ^5^. For regions with multiple conditionally independent pQTLs, we performed pair-wise conditional and colocalization analysis (PWCoCo) ^34^ to obtain colocalization results. PWCoCo conducts conditional analyses to identify independent signals for the two tested traits in a genomic region and then conducts colocalization of each pair of conditionally independent signals for the two traits using summary-level data, which allows for the stringent single-variant assumption to hold for each pair of colocalization analyses.

For the top findings of our MR analysis, we performed colocalization analysis and LD check on single-*cis*-pQTLs with and without full summary data available, respectively, and PWCoCo on multiple-*cis*-pQTLs. For *cis + trans*-pQTLs, we performed colocalization analysis on the *cis* instruments only.

#### MR sensitivity analyses

For multiple-*cis*- and *cis + trans*-pQTL MR top findings, we conducted Cochran’s Q test ^35^ to assess heterogeneity across instrumental SNPs and the MR-Egger intercept test ^36^ to detect unbalanced horizontal pleiotropy. Where the protein did not have an enough number of pQTL instruments in our original pQTL data, we used the data from a recent pQTL study by Gudjonsson et al. ^17^ to conduct these analyses.

To reveal the directionality of effects and avoid reverse causation, we performed Steiger filtering (for all top findings) and bidirectional MR (for those with full summary data available from pQTL studies). Steiger filtering estimates the causal direction by comparing the magnitude of variance explained by the exposure and the outcome ^37^. Bidirectional MR evaluates the evidence for causal effects in the reverse direction by modeling AD as the exposure and plasma proteins as the outcome. Instrumental SNPs for AD were selected based on a threshold of *P* < 5 × 10^-8^ from GWAS after LD clumping (r^2^ < 0.001 and distance located 10000 kb apart from each other). The IVW method ^32^ was used to evaluate the effect of AD on plasma proteins. Where full summary statistics were not available in our original pQTL data, we used full summary data from the pQTL study by Gudjonsson et al. to conduct bidirectional MR analysis.

In addition, we also conducted a single-SNP MR analysis for multiple*-cis*-pQTLs, treating each pQTL as a single instrumental SNP. Results for any novel top-hit proteins (i.e., not included in the main top findings) that survived Bonferroni correction would be picked back to the single-*cis*-pQTL analysis above.

#### Replication

To evaluate the reliability of our proteome-wide MR findings, we repeated MR analysis for the top-hit proteins with the following combinations of exposure-outcome GWASes: 1) Zheng et al. – Schwartzentruber et al., 2) Zheng et al. – Kunkle et al., 3) Gudjonsson et al. – Bellenguez et al., and 4) Zheng et al. – Wightman et al. In each combination, the exposure and outcome GWAS samples were both of European ancestry and did not overlap with each other.

#### Multi-omic validation

We validated our top findings on transcriptomic and epigenomic layers in both blood and brain using summary-data-based Mendelian randomization (SMR) followed by the heterogeneity in dependent instruments (HEIDI) test ^38^. SMR aims to test the potential causal effect of molecular traits on a complex trait of interest using summary data of QTL studies and GWASes. The HEIDI test seeks to address if many SNPs in a single region give Wald ratio estimates that are more different from each other than expected by chance under the assumption that there is a single causal variant, and each SNP only exhibits an effect due to LD with the causal SNP. SMR and the HEIDI test were performed with the above eQTL, sQTL, or mQTL data as the exposure GWAS data and Bellenguez et al.’s AD GWAS data as the outcome GWAS data.

We evaluated multi-omic validation results, along with the previous proteome-wide MR results, using the following criteria:

1. On the proteomic layer, we first selected plasma proteins whose MR results passed the Bonferroni correction for multiple testing (i.e., the top-hit proteins); then, we assessed if these results showed colocalization evidence (i.e., either colocalized or LD-checked).
2. On the transcriptomic layer (i.e., gene expression), we first assessed if any *cis*-eQTL MR results for the genes coding these proteins had a p-value < 0.05 in either blood or brain; if yes, we evaluated if they passed the HEIDI test (i.e., *P*_HEIDI_ > 0.05); if they did, we checked if their effect directions were plausible (i.e., whether the effect of the gene expression level was in the same direction as that of the protein).
3. On the alternative splicing layer, we first assessed if any *cis*-sQTL MR results for the alternative splicing of RNA related to these proteins had a p-value < 0.05 in either blood or brain; if yes, we evaluated if they passed the HEIDI test. The plausibility of effect directions was not considered, as there is no expected direction of the effect of alternative splicing on its downstream protein products.
4. On the epigenomic layer (i.e., DNAm), we first assessed if any *cis*-mQTL MR results for the DNAm in/near the genes coding these proteins had a p-value < 0.05 in either blood or brain; if yes, then we evaluated if they passed the HEIDI test; if they did, then we examined if the CpG sites were located in the promoter area of the gene by manually looking them up on Ensembl (https://www.ensembl.org/); if the CpG sites were in the promoter area, we checked if their effect directions were plausible (i.e., whether the effect of the DNAm level was in the opposite direction to that of the protein).

Given the current availability of QTL data in the multi-omic validation stage and the wide use of targeting drugs ^39^ that provided an opportunity to further explore clinical implications, we followed up on ACE on top of the other top-hit proteins and conducted additional analyses such as multivariable MR for this protein and conventional observational analysis for the medication targeting it.

#### Multivariable Mendelian randomization

Given the important role of ACE (one of the top-hit proteins identified in proteome-wide MR) in blood pressure regulation, we utilized multivariable MR ^40^ to explore if the effect of ACE was dependent on blood pressure. As the pQTL study for ACE used in our main MR analysis did not share full summary data ^41^, we instead employed the pQTL data by Gudjonsson et al. ^17^ used in our replication analysis. To avoid sample overlap, we used the AD GWAS by Kunkle et al. ^20^ as the outcome GWAS, which did not include the UK Biobank samples used for SBP/DBP GWASes. Separate multivariable MR models were applied to SBP and DBP, respectively. SNPs related to SBP/DBP were selected from each GWAS dataset with the *p*-value threshold of 5 × 10^-8^ and combined with the instrumental SNPs for ACE. Genetic association data with ACE and SBP/DBP were extracted for each SNP, followed by deduplication and LD clumping. A total of 244 SBP-related SNPs and 263 DBP-related SNPs, respectively, in addition to the 6 pQTLs for ACE, were included in the multivariable MR analysis. Multivariable MR inverse variance-weighted (MVMR-IVW) estimates ^42^ were computed for each model.

#### Conventional observational study

To further support our MR findings for the protein ACE in the “real-world” setting, we conducted an observational study in the UK Biobank ^43^ to examine the association between the regular use of ACEIs and the risk of AD in later life. Considering potential confounding by indications, regular users of ARBs, another type of antihypertensive with similar indications as ACEIs, were used as the reference group.

Baseline characteristics were presented as frequency (percentage) for categorical variables and mean (SD) for continuous variables. Cox proportional hazards models were applied to examine the longitudinal association between the regular use of ACEIs at baseline and the incidence of AD, with regular use of ARBs as the reference. Three sets of models were performed in the analysis: Model 1 was adjusted for age at recruitment, sex, and ethnicity; Model 2 was additionally adjusted for hypertension, diabetes, hyperlipidemia, traumatic brain injury, cerebrovascular diseases, and nervous system diseases; Model 3 was further adjusted for smoking status, alcohol consumption frequency, BMI, income score, and educational score. Considering that ACEIs and ARBs are primarily used as the treatment of hypertension, we also restricted our analysis to participants with hypertension and repeated the analysis using the aforementioned three models.

In addition, we performed the following sensitivity analyses to test the robustness of our findings: 1) to minimize reverse causality, we excluded the participants who developed AD during the first 2 years of follow-up; 2) considering the potential impact of other chronic diseases, we further adjusted for the number of long-term conditions ^26^; 3) we repeated the analysis after excluding participants with baseline hyperlipidemia and diabetes, respectively.

#### Analysis software

The majority of MR analyses (including Wald ratio, IVW, single-SNP analysis, bidirectional MR, Steiger filtering, and the heterogeneity and pleiotropy tests) were conducted using the “TwoSampleMR” R package ^28^. The IVW analysis considering LD patterns (i.e., generalized IVW) was conducted using the “MendelianRandomization” R package ^31^. The MVMR analysis was conducted by the “MVMR” R package ^44^. Colocalization analysis was conducted by the “coloc” R package ^33^ and the “PWCoCo” tool (https://github.com/jwr-git/pwcoco) ^34^. SMR and HEIDI were performed by the “SMR” software tool (version 1.3.1; downloaded from: https://yanglab.westlake.edu.cn/software/smr/#Download) ^38^. Cox proportional hazards analysis was performed using Stata 17 (Stata Corp. LP, College Station, TX).

#### Data availability

Mendelian randomization and other relevant analyses were based on publicly available data, for example, from publications by Zheng et al. (https://doi.org/10.1038/s41588-020-0682-6), Bellenguez et al. (https://doi.org/10.1038/s41588-022-01024-z), Schwartzentruber et al. (https://doi.org/10.1038/s41588-020-00776-w), Wightman et al. (https://doi.org/10.1038/s41588-021-00921-z), Kunkle et al. (https://doi.org/10.1038/s41588-019-0358-2), and Gudjonsson et al. (https://doi.org/10.1038/s41467-021-27850-z), as well as data resources such as eQTLGen (https://www.eqtlgen.org/), BrainMeta (https://yanglab.westlake.edu.cn/data/SMR/BrainMeta_v1.tar.gz; https://yanglab.westlake.edu.cn/data/SMR/BrainMeta_cis_sqtl_summary.tar.gz), GTEx (https://www.gtexportal.org/), GoDMC (http://mqtldb.godmc.org.uk/), Brain-mMeta (https://yanglab.westlake.edu.cn/data/SMR/Brain-mMeta.tar.gz), and IEU OpenGWAS Project (http://gwas.mrcieu.ac.uk/). Conventional observational analysis corresponds to UK Biobank Project ID 75283. Data from the UK Biobank are available at https://biobank.ndph.ox.ac.uk/ by application. The included GWASes had obtained the necessary ethical approvals from the relevant committees and written informed consent was obtained from all individuals involved in these studies. The UK Biobank study protocol was approved by the North West Multi-centre Research Ethics Committee (11/NW/0382).

## RESULTS

### Proteome-wide MR analysis

After removing proteins with *trans*-only instruments and harmonizing with the summary data from the outcome GWAS data, we obtained 593 single*-cis* pQTL instruments for 593 proteins, 298 multiple*-cis* pQTL instruments for 124 proteins, and 182 *cis + trans* pQTL instruments for 73 proteins. A total of eight [granulin (GRN), angiotensin-converting enzyme (ACE), complement decay-accelerating factor (CD55), triggering receptor expressed on myeloid cells 2 (TREM2), transmembrane protein 106B (TMEM106B), leukocyte immunoglobulin-like receptor B1 (LILRB1), signal-regulatory protein alpha (SIRPA), and sialic acid-binding immunoglobulin-like lectin 9 (SIGLEC9)], five [alpha-L-iduronidase (IDUA), leukocyte immunoglobulin-like receptor B2 (LILRB2), transcobalamin II (TCN2), cathepsin H (CTSH), and glypican 5 (GPC5)], and two [leukocyte immunoglobulin-like receptor A4 (LILRA4) and interleukin 6 signal transducer (IL6ST)] proteins passed the Bonferroni correction (based on the total number of proteins available in each analysis: 593, 124, and 73, respectively) in the MR analysis for single*-cis*, multiple*-cis*, and *cis + trans* pQTL instruments, respectively (**Table 1**). Specifically, among these 15 top-hit proteins, we found evidence [odds ratio (OR) and 95% confidence interval (CI)] of genetically predicted higher plasma GRN [0.79 (0.74-0.84)], ACE [0.91 (0.88-0.93)], CD55 [0.90 (0.86-0.93)], TREM2 [0.67 (0.58-0.78)], LILRB1 [0.95 (0.93-0.97)], TCN2 [0.98 (0.97-0.99)], and LILRA4 [0.89 (0.85-0.95)] levels being associated with lower AD risk, and evidence of genetically predicted higher plasma TMEM106B [1.16 (1.09-1.23)], SIRPA [1.03 (1.02-1.05)], SIGLEC9 [1.03 (1.02-1.04)], IDUA [1.06 (1.05-1.08)], LILRB2 [1.04 (1.02-1.06)], CTSH [1.05 (1.02-1.07)], GPC5 [1.04 (1.02-1.05)], and IL6ST [1.09 (1.04-1.14)] levels being associated with higher AD risk (**Table 1**).

**Table 1.** Top findings from plasma proteome-wide Mendelian randomization.

| Protein | N of SNP | MR Method | OR (95% CI) | Original p-value | Corrected p-value* | Colocalization† | Heterogeneity‡ | Pleiotropy§ | Reverse causation¶ | Bidirectional effect# |
| --- | --- | --- | --- | --- | --- | --- | --- | --- | --- | --- |
| **Single-cis** |  |  |  |  |  |  |  |  |  |  |
| GRN | 1 | Wald ratio | 0.79 (0.74-0.84) | 2.19E-12 | 1.58E-09 | Colocalized | - | - | FALSE | FALSE |
| ACE | 1 | Wald ratio | 0.91 (0.88-0.93) | 6.07E-12 | 4.38E-09 | LD-checked | - | - | FALSE | - |
| CD55 | 1 | Wald ratio | 0.90 (0.86-0.93) | 1.62E-09 | 1.17E-06 | LD-checked | - | - | FALSE | FALSE |
| TREM2 | 1 | Wald ratio | 0.67 (0.58-0.78) | 6.95E-08 | 5.02E-05 | LD-checked | - | - | FALSE | - |
| TMEM106B | 1 | Wald ratio | 1.16 (1.09-1.23) | 1.92E-06 | 1.38E-03 | LD-checked | - | - | FALSE | - |
| LILRB1 | 1 | Wald ratio | 0.95 (0.93-0.97) | 4.04E-06 | 2.92E-03 | LD-checked | - | - | FALSE | FALSE |
| SIRPA | 1 | Wald ratio | 1.03 (1.02-1.05) | 1.37E-05 | 9.88E-03 | LD-checked | - | - | FALSE | - |
| SIGLEC9 | 1 | Wald ratio | 1.03 (1.02-1.04) | 4.30E-05 | 3.10E-02 | Not colocalized | - | - | FALSE | FALSE |
| **Multiple-cis** |  |  |  |  |  |  |  |  |  |  |
| IDUA | 3 | Generalized IVW | 1.06 (1.05-1.08) | 6.37E-21 | 4.60E-18 | Colocalized | TRUE | FALSE | FALSE | FALSE |
| LILRB2 | 3 | Generalized IVW | 1.04 (1.02-1.06) | 1.37E-06 | 9.90E-04 | Not colocalized | TRUE | FALSE | FALSE | FALSE |
| TCN2 | 4 | Generalized IVW | 0.98 (0.97-0.99) | 2.87E-06 | 2.07E-03 | Not colocalized | FALSE | FALSE | FALSE | FALSE |
| CTSH | 3 | Generalized IVW | 1.05 (1.02-1.07) | 2.00E-05 | 1.44E-02 | Not colocalized | FALSE | FALSE | FALSE | FALSE |
| GPC5 | 3 | Generalized IVW | 1.04 (1.02-1.05) | 3.24E-05 | 2.34E-02 | Not colocalized | FALSE | FALSE | FALSE | FALSE |
| **cis + trans** |  |  |  |  |  |  |  |  |  |  |
| LILRA4 | 2 (1 cis) | IVW | 0.89 (0.85-0.95) | 9.05E-05 | 6.61E-03 | Not colocalized | FALSE | - | FALSE | FALSE |
| IL6ST | 4 (1 cis) | IVW | 1.09 (1.04-1.14) | 1.51E-04 | 1.10E-02 | Not colocalized | FALSE | FALSE | FALSE | FALSE |

\* Corrected using the Bonferroni method (single-cis: 593 proteins; multiple-cis: 124 proteins; cis + trans: 73 proteins).
† Findings with evidence for colocalization (i.e., posterior probability for a shared causal variant > 80%) shown as “Colocalized”; findings without evidence for colocalization but passing LD-check shown as “LD-checked”; others shown as “Not colocalized”; colocalization analysis not applicable for some findings due to the unavailability of full summary data.
‡ Tested by Cochran’s Q statistics, with p-value < 0.05 defined as having evidence for heterogeneity (shown as “TRUE”); not applicable for some findings due to insufficient numbers of SNPs.
§ Tested by the MR-Egger intercept test, with p-value < 0.05 defined as having evidence for unbalanced horizontal pleiotropy (shown as “TRUE”); not applicable for some findings due to insufficient numbers of SNPs.
¶ Tested by Steiger filtering, with p-value < 0.05 defined as having evidence for reverse causation (shown as “TRUE”).

### Colocalization analysis

Among the eight proteins with single-*cis* pQTLs (four with full summary data available), only GRN showed colocalization evidence (PP for a shared causal variant = 99.4%, **Table 1**, **Figure S1**). In the absence of full summary data or colocalization evidence detected by colocalization analysis, LD check revealed evidence for approximate colocalization for ACE, CD55, TREM2, TMEM106B, LILRB1, and SIRPA (**Table 1**). For the five proteins with multiple-*cis* pQTLs, PWCoCo showed colocalization evidence for IDUA (**Table 1**). For the two proteins with *cis* + *trans* pQTLs, colocalization analysis (on the *cis*-pQTL only) failed to find evidence for colocalization in either protein. See regional plots of all proteins with full GWAS summary statistics in **Figure S1-S11**.

When using the data from a recent pQTL study by Gudjonsson et al.,^17^ where full summary statistics were available for ACE, TREM2, TMEM106B, and SIRPA, colocalization evidence was found for ACE and TMEM106B (**Table S1**).

### Multi-omics validation and selection of top findings

Detailed SMR and HEIDI results are shown in **Table S2-S7**. Of the 15 top-hit plasma proteins that passed the Bonferroni correction threshold, GRN, ACE, CD55, TREM2, TMEM106B, LILRB, SIRPA, IDUA, and TCN2 showed evidence for colocalization (including approximate colocalization detected by LD-check); in the blood/brain *cis-*eQTL SMR analysis, the expression of three genes (*ACE*, *SIGLEC9*, and *IDUA*) was associated with AD risk at the *P* = 0.05 level, passed the HEIDI test (i.e., *P*_HEIDI_ > 0.05), and had plausible effect directions (i.e., in the same direction as the pQTL MR results); in the blood/brain *cis-*sQTL SMR analysis, alternative splicing in five genes (*ACE*, *TMEM106B*, *SIRPA*, *SIGLEC9*, and *IDUA*) was associated with AD risk at the *P* = 0.05 level and passed the HEIDI test; in the blood/brain *cis-*mQTL SMR analysis, we only found that DNAm in the *ACE* gene was associated with AD risk at the *P* = 0.05 level, passed the HEIDI test, was located in the promoter area of the gene (cg06751221, 17:63477520-63477569), and had a plausible effect direction (i.e., in the opposite direction as the pQTL MR results) (**Figure 2**, **Table 2**; see detailed sQTL MR results in **Figure S12-S13**). It should also be noted that validation analyses were not always applicable due to the unavailability of QTL data for some proteins.

**Figure 2.**
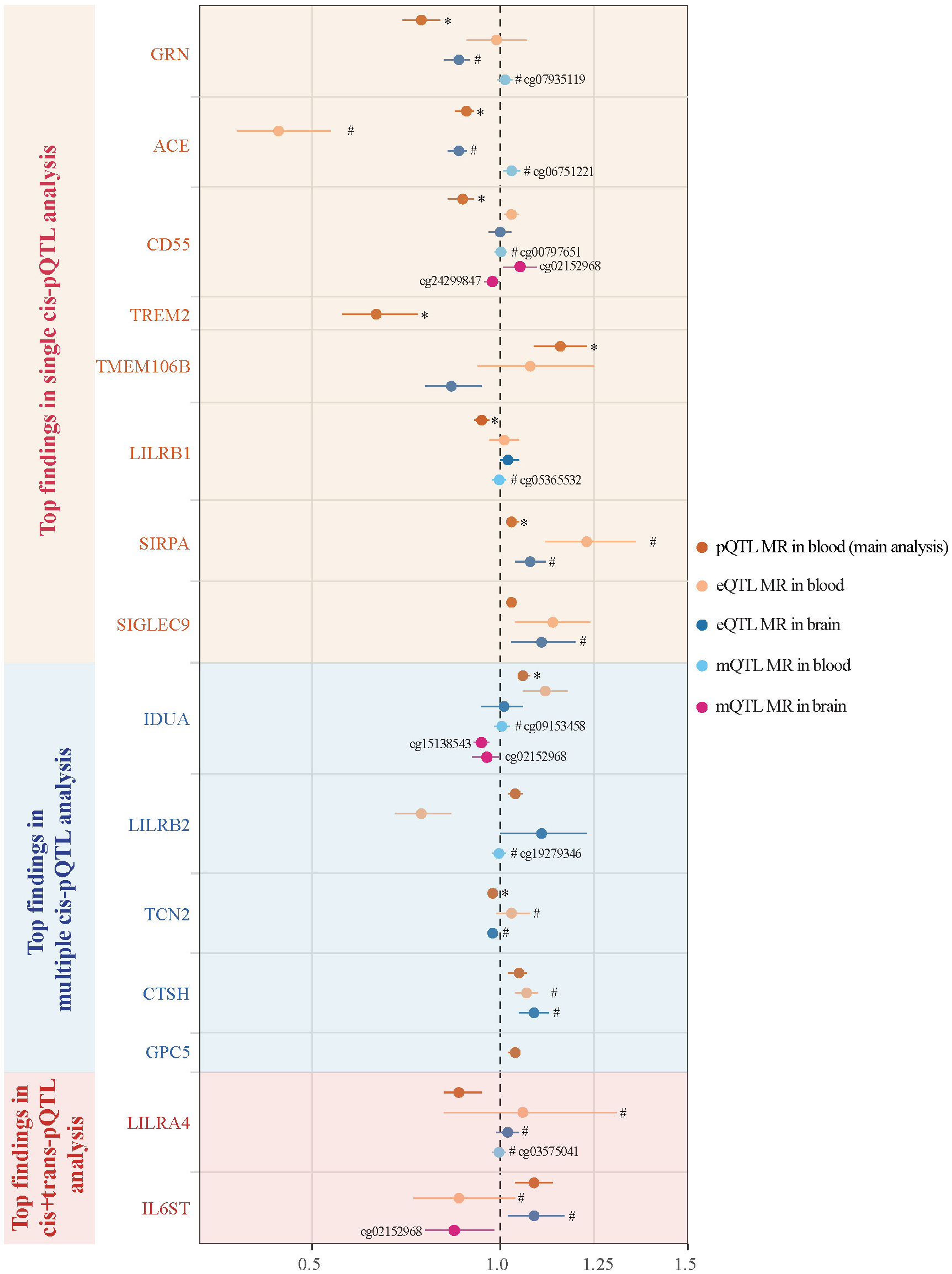
Multi-omic validation results in blood and brain for the preliminary top findings. Only eQTL and mQTL MR results are shown here; see detailed sQTL MR results in **Figure S12-13**. * With evidence for colocalization (i.e., either colocalized or LD-checked). # Passed the HEIDI test (i.e., *P*_HEIDI_ > 0.05). pQTL, protein quantitative trait locus. MR, Mendelian randomization. eQTL, expression quantitative trait locus. mQTL, methylation quantitative trait locus. sQTL, splicing quantitative trait locus. LD, linkage disequilibrium. HEIDI, heterogeneity in dependent instruments.

**Table 2.**
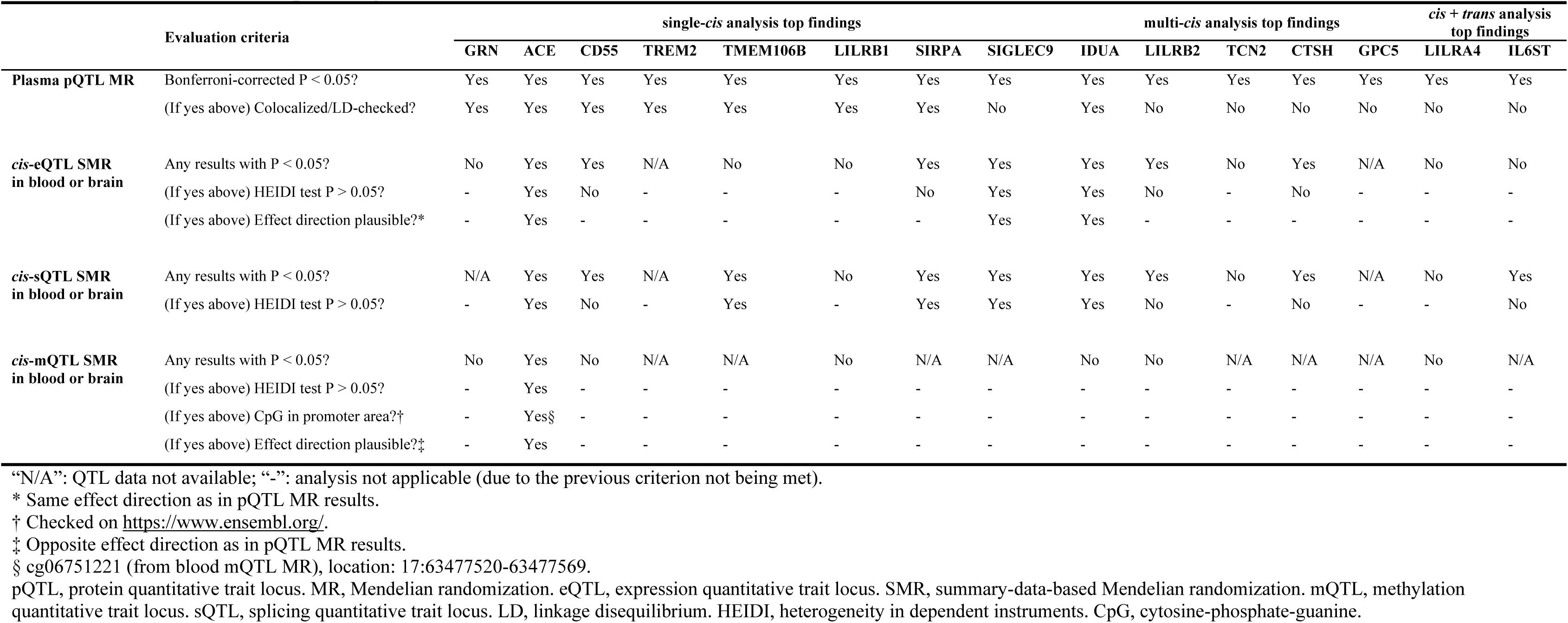
Evaluation of the top findings.

|  | Evaluation criteria | single-*cis* analysis top findings |  |  |  |  |  |  |  | multi-*cis* analysis top findings |  |  |  |  | *cis* + *trans* analysis top findings |  |
| --- | --- | --- | --- | --- | --- | --- | --- | --- | --- | --- | --- | --- | --- | --- | --- | --- |
|  |  | GRN | ACE | CD55 | TREM2 | TMEM106B | LILRB1 | SIRPA | SIGLEC9 | IDUA | LILRB2 | TCN2 | CTSH | GPC5 | LILRA4 | IL6ST |
| Plasma pQTL MR | Bonferroni-corrected P < 0.05? | Yes | Yes | Yes | Yes | Yes | Yes | Yes | Yes | Yes | Yes | Yes | Yes | Yes | Yes | Yes |
|  | (If yes above) Colocalized/LD-checked? | Yes | Yes | Yes | Yes | Yes | Yes | Yes | No | Yes | No | No | No | No | No | No |
| *cis*-eQTL SMR in blood or brain | Any results with P < 0.05? | No | Yes | Yes | N/A | No | No | Yes | Yes | Yes | Yes | No | Yes | N/A | No | No |
|  | (If yes above) HEIDI test P > 0.05? | - | Yes | No | - | - | - | No | Yes | Yes | No | - | No | - | - | - |
|  | (If yes above) Effect direction plausible?* | - | Yes | - | - | - | - | - | Yes | Yes | - | - | - | - | - | - |
| *cis*-sQTL SMR in blood or brain | Any results with P < 0.05? | N/A | Yes | Yes | N/A | Yes | No | Yes | Yes | Yes | Yes | No | Yes | N/A | No | Yes |
|  | (If yes above) HEIDI test P > 0.05? | - | Yes | No | - | Yes | - | Yes | Yes | Yes | No | - | No | - | - | No |
| *cis*-mQTL SMR in blood or brain | Any results with P < 0.05? | No | Yes | No | N/A | N/A | No | N/A | N/A | No | No | N/A | N/A | N/A | No | N/A |
|  | (If yes above) HEIDI test P > 0.05? | - | Yes | - | - | - | - | - | - | - | - | - | - | - | - | - |
|  | (If yes above) CpG in promoter area?† | - | Yes§ | - | - | - | - | - | - | - | - | - | - | - | - | - |
|  | (If yes above) Effect direction plausible?‡ | - | Yes | - | - | - | - | - | - | - | - | - | - | - | - | - |

“N/A”: QTL data not available; “-”: analysis not applicable (due to the previous criterion not being met).
\* Same effect direction as in pQTL MR results.
† Checked on https://www.ensembl.org/.
‡ Opposite effect direction as in pQTL MR results.
§ cg06751221 (from blood mQTL MR), location: 17:63477520-63477569.
pQTL, protein quantitative trait locus. MR, Mendelian randomization. eQTL, expression quantitative trait locus. SMR, summary-data-based Mendelian randomization. mQTL, methylation quantitative trait locus. sQTL, splicing quantitative trait locus. LD, linkage disequilibrium. HEIDI, heterogeneity in dependent instruments. CpG, cytosine-phosphate-guanine.

Given the current availability of QTL data in the multi-omic validation stage and the wide use of targeting drugs ^39^ that provided an opportunity to further explore clinical implications, we followed up on ACE (angiotensin-converting enzyme) on top of the other 14 top-hit proteins and conducted additional analyses such as multivariable MR for this protein and observational analysis for the medication targeting it.

### MR sensitivity analyses

We conducted a range of sensitivity and replication analyses to test the robustness and reliability of our MR findings. For ACE specifically (see other proteins in **Text S1**), our sensitivity analyses did not find evidence for reverse causation as detected by Steiger filtering, whereas Cochran’s Q test, MR-Egger intercept test, and bidirectional MR were not applicable as ACE had only one pQTL instrument and its GWAS full summary statistics were not available. Where these sensitivity analyses were not applicable due to the insufficient number of instruments or the unavailability of full summary data, we used the data from a recent pQTL study by Gudjonsson et al. (where multiple pQTL instruments and full summary statistics were available) and found no evidence for heterogeneity (by Cochran’s Q test), pleiotropy (by MR-Egger intercept test), or reverse causation (by bidirectional MR) for ACE (**Table S1**). See the detailed description of sensitivity analysis results (including single-SNP analysis results in **Table S8**) for other top-hit proteins in **Text S1**.

In replication analyses, we used data from different pQTL studies and GWASes to replicate our MR top findings. We first used an earlier AD GWAS by Schwartzentruber et al. [0.93 (0.90-0.96)] and Wightman et al. [0.987 (0.981-0.992)] and then a smaller but more clinically relevant AD GWAS by Kunkle et al. (only included clinically diagnosed AD cases) [0.92 (0.88-0.97)] as the outcome GWAS and observed similar results in effect directions and sizes for ACE (**Table S9**, **Figure S14**). Then, we replaced our original pQTL data with those from a recently published GWAS of 2,091 serum proteins by Gudjonsson et al. and also found that ACE had a similar effect estimate [0.93 (0.90-0.97)] as our main MR results (**Table S9**, **Figure S14**). See the detailed description of replication analysis results for other top-hit proteins in **Text S1**.

Our multivariable MR showed that after adjusting for SBP or DBP, the effect estimate of ACE did not differ from our univariable (i.e., main) MR results (**Table S10**, **Figure S15**), suggesting that the protective effect of ACE on AD risk was likely independent of blood pressure.

### Observational analysis of ACE inhibitor use

From the pQTL MR analysis above, we found a protective effect of higher plasma ACE levels on AD risk [OR (95% CI): 0.91 (0.88-0.93)], suggesting that ACE inhibition (i.e., inhibiting the effect of ACE) might be potentially associated with an increased risk of AD [OR (95% CI): 1.10 (1.08-1.14), estimated by taking the inverse of the ACE effect]. To follow up on this finding and explore its clinical relevance, we used “real-world” drug use data of ACE inhibitors (ACEIs, with ACE inhibition effects) from the UK Biobank and examined if ACEI use was associated with increased AD risk. ACEIs are a commonly used antihypertensive in clinical practice and the only existing class of drugs targeting ACE. To avoid potential confounding by indications, we used angiotensin receptor blockers (ARBs), which have similar indications but different drug targets as ACEIs, as the reference for the comparison.

A total of 59,117 UK Biobank participants were included in the study, including 42,944 ACEI users and 16,173 ARB users. At baseline, the mean age was 60.4 (standard deviation 6.6) years [60.2 (6.7) years in ACEI users vs. 61.0 (6.4) years in ARB users], and 41% of the participants were female (38.3% in ACEI users vs. 48.3% in ARB users; see **Table S11**). Over 723,142 person-years of follow-up [median (interquartile range) length of follow-up: 12.77 (11.92–13.54) years], there were 552 cases of AD, with 0.97% (416/42,944) in ACEI users and 0.84% (136/16,173) in ARB users.

Analysis with Cox proportional hazard models showed a higher incidence of AD in ACEI users compared with ARB users [hazard ratio (95% CI): 1.24 (1.01-1.52)] (**Figure 3**), with adjustment for age at recruitment, sex, ethnicity, pre-existing morbidities (hypertension, diabetes, hyperlipidemia, cerebrovascular diseases, traumatic brain injury, and diseases of the nervous system), smoking status, alcohol use, body mass index, income, and educational.

**Figure 3.**
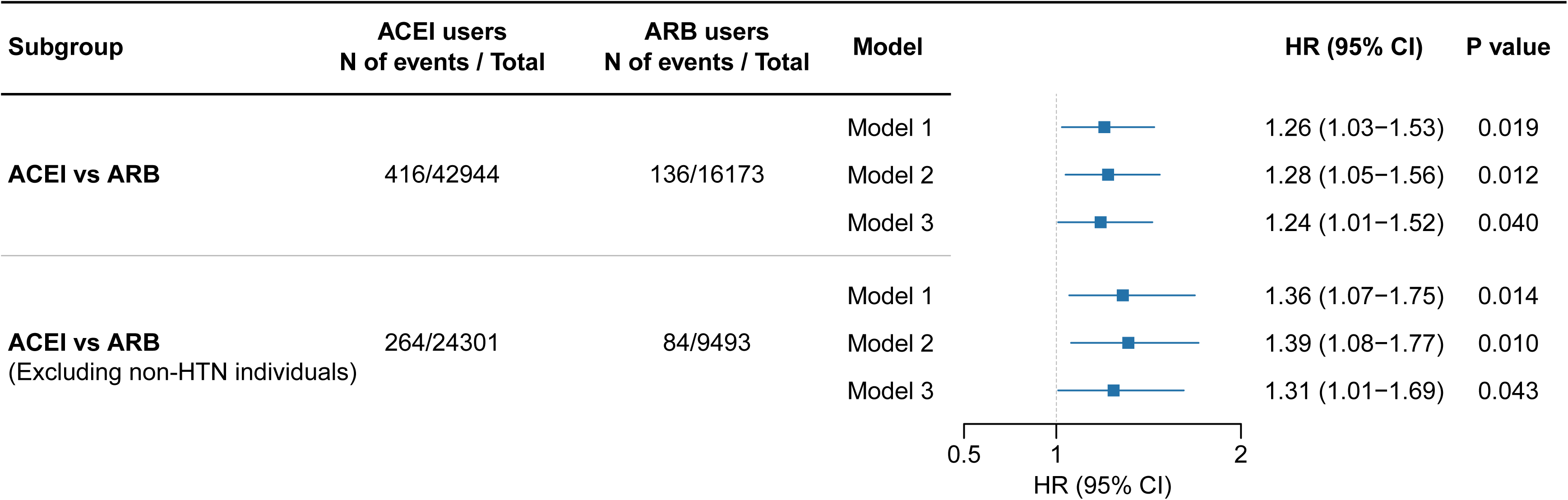
Association of angiotensin-converting enzyme inhibitor vs. angiotensin receptor blocker with the incidence of Alzheimer’s disease in UK Biobank. Model 1 was adjusted for age at recruitment, sex, and ethnicity. Model 2 was additionally adjusted for hypertension, diabetes, hyperlipidemia, cerebrovascular diseases, traumatic brain injury, and diseases of the nervous system. Model 3 was further adjusted for smoking status, alcohol use, body mass index, income, and educational level. ACEI, angiotensin-converting enzyme inhibitor. ARB, angiotensin receptor blocker. HTN, hypertension. HR, hazard ratio. CI, confidence interval.

When restricting to participants with hypertension, the results were consistent [1.31 (1.01-1.69)] (**Figure 3**). Moreover, no major changes in the results were found in sensitivity analyses excluding cases with baseline diabetes or hyperlipidemia, additionally adjusting for the number of baseline chronic conditions, or removing AD cases occurring within the first two years of follow-up (**Figure S16**).

Given the MR evidence suggesting that inhibition of ACE might increase the risk of AD, the higher AD risk associated with the regular use of ACEIs (which inhibit ACE activity) vs. their counterpart ARBs (block angiotensin II receptors) observed in the UK Biobank complements our MR findings and provides further evidence for the implications of these findings in clinical settings (**Figure 4**).

**Figure 4.**
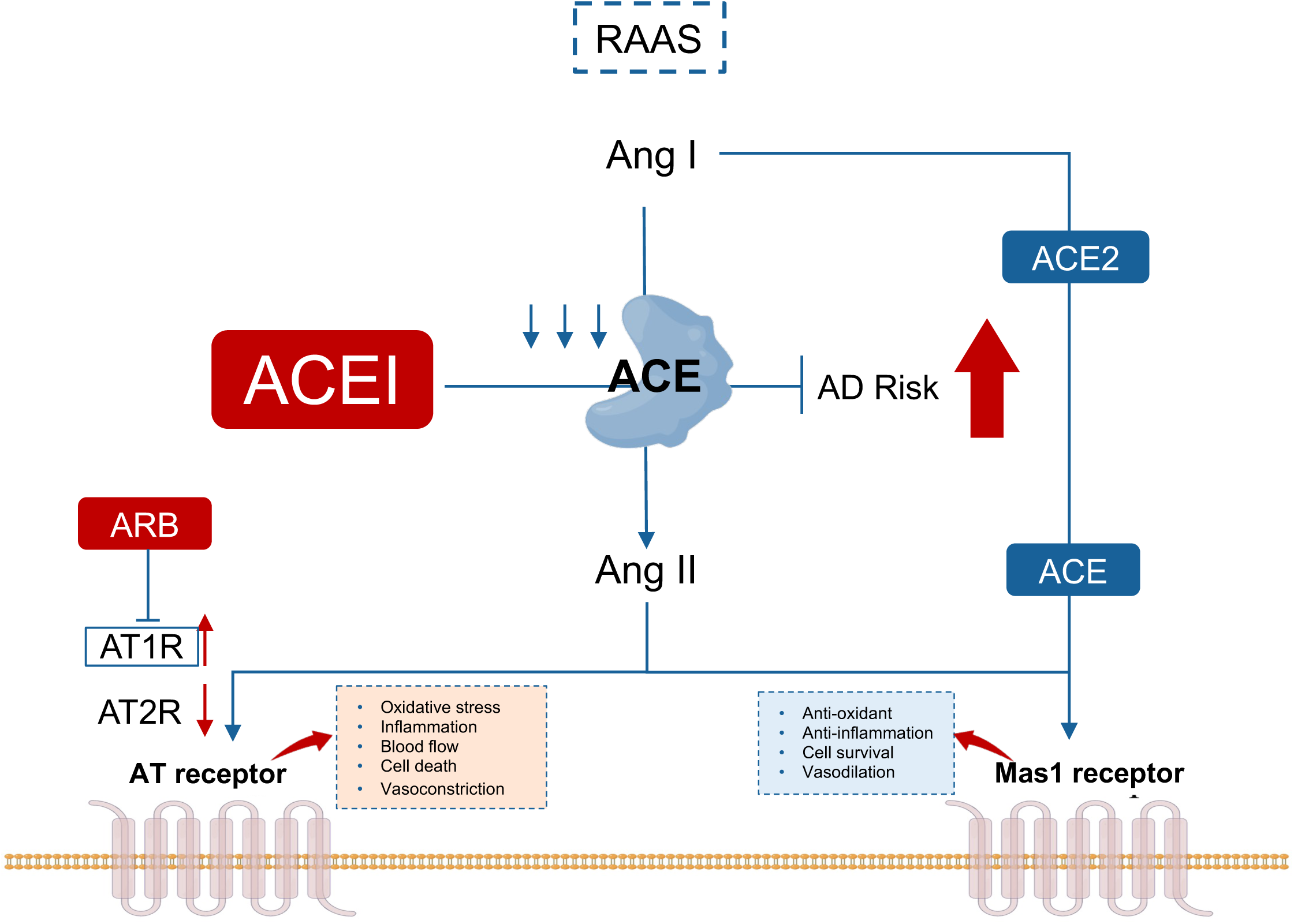
The use of ACE inhibitors is associated with AD risk as compared with ARBs. RAAS, renin-angiotensin-aldosterone system; Ang I/II, Angiotensin I/II; ACE, angiotensin-converting enzyme; ARB, angiotensin II receptor blockers; AT receptor, Angiotensin receptor. AD, Alzheimer’s disease.

## DISCUSSION

In this study, we conducted a proteome-wide MR analysis, which identified 15 plasma proteins associated with AD risk, including ACE, GRN, CD55, TREM2, TMEM106B, LILRB1, SIRPA, SIGLEC9, IDUA, LILRB2, TCN2, CTSH, GPC5, LILRA4, and IL6ST, followed by validation on the transcriptomic and epigenomic layers. Of these proteins, inhibition of ACE was suggested to increase the risk of AD, which was likely independent of blood pressure. Given the availability of QTL data in the multi-omic validation stage and the popularity of relevant drugs, we further explored the clinical implications of ACE and its targeting drugs. As a classic antihypertensive medication inhibiting ACE activity in “real-world” settings, ACEIs were observationally associated with an increased risk of AD as compared with ARBs, another antihypertensive with similar indications but different targets, in the UK Biobank, which complements our MR results.

Our findings generally align with and expand upon prior research on proteins as potential druggable targets for preventing AD. For example, a recent study by Ou et al. applied similar methodologies (e.g., proteome-wide MR, colocalization, and eQTL MR) to explore potential AD drug targets by integrating genetics and proteomes from the brain and blood ^6^. They identified seven genes (*ACE*, *ICA1L*, *TOM1L2*, *SNX32*, *EPHX2*, *CTSH*, and *RTFDC1*) with brain protein abundance causally linked to AD. Among these, the protective effect of *ACE* was substantiated by both proteomic and transcriptomic evidence in blood. Our study, which yielded comparable results for ACE, provides robust support for their findings by discovering consistent evidence supporting our MR findings for ACE from observational analysis of “real-world” drug use data, addressing the methodological limitations of using a single epidemiologic approach ^45^ and underscoring the implications of our findings for clinical practice. Moreover, we also leveraged larger and more recent GWAS data, verified the reliability of findings across multiple pQTL studies and AD GWASes, and further validated the findings at the alternative splicing and DNA methylation levels. Additionally, despite not having brain pQTL data, our study still supports their finding for CTSH by providing stronger evidence in blood. To sum up, Ou et al.’s and our studies serve as complementary pieces, collectively contributing to drug discovery, repurposing, and vigilance efforts for AD.

Specifically, the protective effect of the protein ACE against AD is a noteworthy finding in both Ou et al.’s and our studies that warrants further discussion. While previous genetic and epidemiologic studies have reported similar associations ^6, 46–48^, our work provides deeper insights into the potential mechanisms underlying the relationship between ACE and AD risk. ACE is well-known for its biological functions, such as its involvement in the renin-angiotensin system (RAS), which plays a crucial role in regulating blood pressure ^49^.

However, our multivariable MR results suggest that the effect of ACE on AD risk seemed to be independent of blood pressure. In our observational analysis of prospective cohort data, a difference in AD risk was also observed between users of ACEIs and ARBs, which both target the RAS but act on different molecules (ACEIs inhibit the angiotensin-converting enzyme, while ARBs block angiotensin II receptors, as shown in **Figure 4**) ^50^. All these findings suggest that ACE may influence neurodegenerative diseases through unique pathways other than blood pressure regulation ^51^. One such potential mechanism is the degradation of amyloid-beta (Aβ) deposits. ACE is a component of Aβ-degrading enzymes (ADEs) and can facilitate the clearance of Aβ deposits, which are implicated in the pathogenesis of AD ^52^.

The discovery that ACEI use was associated with increased AD risk raises important questions for clinical and public health practice. ACEIs and ARBs are both widely used first-line antihypertensives, which are usually recommended interchangeably by current guidelines ^53^. In the United States alone, 13.5 million people take ARBs and 19.1 million take ACEIs each year ^54^. Although only a 20-30% increased AD risk associated with ACEI use was observed in our study, considering the huge number of ACEI users and the increasingly heavy burden of AD ^1^, its public health implications are substantial on a global scale. If our findings can be confirmed by high-quality clinical trials, switching first-line RAS blockers from ACEIs to ARBs may have a great impact on population health. Preferentially prescribing ARBs over ACEIs is also supported by a recent statement by Messerli et al. ^39^ and a large study ^55^ of 2,297,881 patients initiating treatment with ACEIs and 673,938 patients with ARBs, which has found that while ACEIs and ARBs have similar efficacy in reducing the risk of cardiovascular events, ARB users have better safety outcomes such as lower risks of angioedema, cough, and gastrointestinal conditions. In addition, further research is needed to elucidate the complex relationship between ACE, blood pressure, other antihypertensive medications, and AD risk, and to determine the potential impact of these findings on the management of hypertension and prevention strategies for AD. Our findings highlight the need to carefully consider the potential risks and benefits of ACEIs and other antihypertensive medications in the context of neurodegenerative diseases.

Several other proteins identified in our study have been previously linked to the pathogenesis of AD or other neurodegenerative diseases by in-vivo and in-vitro studies. For example, granulin (GRN) and triggering receptor expressed on myeloid cells 2 (TREM2) have been shown to play a role in microglial function and neuroinflammation ^56–58^, which are key processes in AD development ^59^. Other identified proteins, such as CD55 and TMEM106B, have also been reported to have potential roles in AD-related pathways, including immune response and cellular homeostasis. The protein CD55 has been shown to mitigate neuronal cell death and apoptosis during hypoxia induced by sodium cyanide ^60^. A study of aged mice with *Tmem106b* knockout, heterozygote, and wild-type genotypes found that TMEM106B-dependent lysosomal trafficking defects led to neuronal dysfunction and behavioral impairments ^61^. Therefore, our findings for other top-hit proteins warrant further investigations into their mechanistic contributions to AD and their potential value as drug targets for AD prevention.

Our study has several strengths, one of which is the application of a proteome-wide MR approach. This method enabled us to systematically evaluate the effects of plasma proteins on AD risk while minimizing the influence of confounding and reverse causation. By integrating multiple omic layers, including proteomic, transcriptomic, epigenomic, and genomic data, we provided comprehensive evidence to support the reliability of our results. Importantly, we also combined MR with observational analysis of prospective cohort data, establishing a connection between molecule-level research and “real-world” clinical evidence, which strengthens the clinical relevance of our findings.

Despite these strengths, our study also has the following limitations. First, although MR is a powerful approach for addressing confounding, it cannot eliminate the possibility of horizontal pleiotropy ^62^, and the MR methods we used could only examine the linear relationship between the protein level and AD risk. Second, our study only assessed proteins with available genetic instruments of relatively high reliability (i.e., tier 1 instruments in Zheng et al.’s study) ^5^, which may have excluded other potentially relevant proteins in the context of AD pathogenesis. Similarly, an additional limitation is the absence of QTL data (e.g., mQTL data in blood and brain) for some top-hit proteins, which precluded our follow-up analyses on many potentially relevant results. Fourth, as our study predominantly included participants of European descent, the generalizability of our findings to other populations remains uncertain. It is essential to validate our results in diverse ethnic groups to better understand the applicability of our findings across different populations ^63^. Another limitation is the potential of residual confounding in our observational analysis due to unmeasured or inaccurately measured confounders, although the consistency between observational and MR evidence mitigates this concern to some extent. In addition, our observational analysis with baseline drug use as the exposure (i.e., prevalent users) may be subject to selection bias, and future research can explore the effect of initiation, maintenance, or discontinuation of ACEI use on AD risk by explicitly emulating a target trial using observational data ^64^. Selection bias might also occur when restricting our observational analysis to ACEI and ARB users, although this restriction was justified to avoid confounding by indication. Lastly, while we were able to identify some proteins with possible causal effects on AD risk, it is important to recognize that these proteins may not be acting independently. The complex interplay between these proteins and their roles in various biological pathways could contribute to the observed associations, warranting further investigation to fully elucidate the intricate network of molecular mechanisms underlying AD pathogenesis.

In conclusion, our proteome-wide MR study identified 15 plasma proteins associated with AD risk. Of these, ACE inhibition was suggested to increase the risk of AD. Observationally, the regular use of ACEIs, a common class of antihypertensive inhibiting the effect of ACE, was associated with an increased risk of AD as compared with ARBs, another antihypertensive with similar indications but different molecular targets. While this key finding warrants further investigation, it underscores the need to carefully consider potential implications in terms of neurodegenerative diseases when prescribing ACEIs for high-risk populations and add support to the preferential prescription of ARBs over ACEIs in clinical practice. Our work also highlights the clinical translatability of interdisciplinary research integrating genetic/omics-based approaches with “real-world” data in the field of pharmaco-epidemiology.

## ACKNOWLEDGMENTS

We thank all participants and researchers from GWAS consortia, QTL databases, and the UK Biobank.

## SOURCES OF FUNDING

This work was supported by grants from the National Key Research and Development Program of China (2022YFC2505203, 2022YFC3602400, 2022YFC3602401), National Natural Science Foundation of China (82170437), Central South University Innovation-Driven Research Program (2023CXQD007). JZ is a member of the Innovative Research Team of High-level Local Universities in Shanghai. TRG and ES work at a unit that receives funding from the University of Bristol and the UK Medical Research Council (MC_UU_00011/1, MC_UU_00032/01, MC_UU_00032/03 and MC_UU_00011/4). T.R.G. also holds a Turing Fellowship from the Alan Turing Institute. PH is funded by Wellcome Trust PhD Studentship in Molecular, Genetic and Lifecourse Epidemiology (224979/Z/22/Z). The funding sources were not involved in the study design, the interpretation of data, the writing of the report, and the decision to submit the article for publication.

## DISCLOSURES

Tom R Gaunt declares that he receives funding from Biogen for other research not represented in this manuscript, while other authors declare no conflict of interest.

## Notes

### Clinical Trial

This study is not a clinical trial.

### Author Declarations

Conventional observational analysis corresponds to UK Biobank Project ID 75283. Data from the UK Biobank are available at biobank.ndph.ox.ac.uk/ by application. The included GWASes had obtained the necessary ethical approvals from the relevant committees and written informed consent was obtained from all individuals involved in these studies. The UK Biobank study protocol was approved by the North West Multi-centre Research Ethics Committee (11/NW/0382).

